# Acupuncture at Sanyinjiao(SP6) for patients with primary dysmenorrhea: Protocol for a systematic review and meta-analysis

**DOI:** 10.1101/2023.09.24.23296053

**Authors:** Xuemei Wang, Jiao Yang, Jing Liu, Yepeng Yang, Zhiyuan Zhang, Ping Xie

**Affiliations:** Department of Gynecology, Hospital of Chengdu University of Traditional Chinese Medicine; College of Clinical Medicine, Chengdu University of Traditional Chinese Medicine, Chengdu, Sichuan Province, P. R. China; West China Second University Hospital, Sichuan University / West China women’s and children’s Hospital; Shanxi University of Chinese Medicine; Dujiangyan Traditional Chinese Medicine Hospital, Chengdu, Sichuan Province, P.R. China

**Keywords:** acupuncture, sanyinjiao, SP6, primary dysmenorrhea, systematic review protocol

## Abstract

**Background:** Primary dysmenorrhea (PD) is a gynecological disease that seriously affects women’s physical and mental health and quality of life. Sanyinjiao (SP6) is the most used acupoint for acupuncture treatment of PD, and acupuncture SP6 is more commonly used in the clinical treatment of PD, but evidence of effectiveness and safety is lacking. The purpose of this systematic review plan is to develop a study protocol that can evaluate the efficacy and safety of acupuncture SP6 in the treatment of PD.

**Methods:** The upper limit of retrieval time will be set to September 2023. Foreign and Chinese databases will be searched respectively: Cochrane Library, PubMed, EMBASE, Ovid, Korea Med, J-Global, J-Stage databases, and CNKI, CBM, SinoMed, VIP, Wanfang. Randomized controlled trials meeting the inclusion criteria will be screened by two independent reviewers, and data extraction and risk of bias assessment will be carried out. The primary outcome is dysmenorrhea symptoms, and the secondary outcomes include recent clinical cure rate, concomitant symptoms, serum prostaglandins, uterine artery blood flow resistance under ultrasound monitoring, and adverse reactions. Use RevMan V.5.3 software to perform the following operations: data synthesis, subgroup analysis, heterogeneity analysis, and sensitivity analysis. The evidence quality of clinical studies included in this study will be evaluated by the software of “Grading of Recommendations Assessment, Development, and Evaluation”.

**Results:** This study will provide comprehensive clinical evidence for treating PD with acupuncture SP6.

**Conclusion:** This systematic review will confirm whether acupuncture SP6 is safe and effective in treating PD clinically.

**Registration details:** PROSPERO CRD42020171664.

## Background

Primary dysmenorrhea (PD) manifests as the main symptoms of periodic lower abdominal pain in women during or after menstruation, accompanied by other discomforts, such as cold sweat, dizziness, nausea, vomiting, diarrhea, fatigue, palpitation, etc., to affect work and life, and gynecological examination indicates that there is no obvious organic disease in reproductive organs [1]. PD, a common clinical gynecological disease, is most common in young unmarried women and has a significant impact on women’s health, life and work. It is also the most common cause of absenteeism among women [2, 3]. PD is a huge public health burden. The prevalence of PD reported abroad ranges from 34% to 94%, among which 61% are moderate dysmenorrhea [4]. A survey of 2000 female adolescents shows that the prevalence of dysmenorrhea is 84.8 % (n = 1080), the average pain score is 5.87, about 34% (n = 439) of women have taken painkillers, and the average absenteeism rate is 18%[5].

At present, the pathogenesis of primary dysmenorrhea is not completely clear. It is generally believed that it is directly related to the increase of prostaglandin, and it is also related to the secretion and synthesis of vasopressin[6], β-endorphin[7], interleukin[8], oxytocin[9], estrogen, and progesterone[10, 11]. Its comprehensive effect causes pathological contraction of uterine smooth muscle, resulting in uterine ischemia and hypoxia, resulting in dysmenorrhea [12, 13]. The commonly used drugs for clinical treatment of PD are non-steroidal anti-inflammatory drugs (NSAIDs), antispasmodic drugs, sedatives, contraceptive-based, the effect is exact [14]. However, these drugs can only temporarily relieve pain, are not recommended for repeated use, and have disadvantages such as limited efficiency and side effects. It is impossible to carry out treatment for patients with no or low response to such drugs, treatment contraindications, treatment intolerance, or non-acceptance.

Acupuncture is a common method of treating PD with traditional Chinese medicine (TCM), of which Sanyinjiao (SP6) is the most used acupoint. Acupuncture SP6 is widely used in the treatment of PD and is one of the alternative methods for the treatment of PD[15 -17]. In addition, acupuncture has almost no side effects such as dizziness, headache, lethargy, gastrointestinal bleeding, and drug dependence. There is some mechanism of acupuncture SP6.

Acupoint SP6 is located on the inside of the ankle joint of human lower limbs and is the intersection of the meridians of the liver, spleen, and kidney. According to the theory of TCM, the main pathogenesis of PD is the imbalance of Chongren and blocked meridians. Acupuncture SP6 has the functions of regulating Chongren, dredging collaterals, and relieving pain. Although the mechanism of acupuncture SP6 in the treatment of PD has not been fully elucidated. However, animal experiments show that acupuncture SP6 can play a role through the following mechanisms [18]: (1) improving uterine microcirculation disturbance; (2) regulate the endocrine function; (3) improve the function of the immune system; (4) affect the metabolism of nerve and neurotransmitters.

In 2013, a systematic review of acupuncture SP6 for PD, the experimental group and control group were acupuncture SP6 and GB39, respectively. The outcome index was only related to the visual analog scale (VAS) pain score. This study failed to evaluate the efficacy, improvement of accompanying symptoms, adverse reactions, and safety of acupuncture SP6 for PD. Acupuncture is widely used in East Asia, represented by China, Japan, and Korea. However, in this study, only four English databases were retrieved, not East Asian databases. Therefore, there are some imperfections in the design of the systematic review, which may affect the authenticity of its research results. In recent years, the clinical research of acupuncture SP6 in the treatment of PD has gradually increased, and the efficacy evaluation indicators have also gradually increased [15, 19, 20]. Therefore, the aim of this systematic review is to evaluate the efficacy and safety of acupuncture at SP6 in the treatment of PD. This work will provide reliable clinical evidence with objective results by formulating strict retrieval strategies and standardized research programs.

## Methods

### Registration and ethics

This systematic review protocol has been registered on PROSPERO international platform with the registration number CRD 42020171664 (https://www.crd.york.ac.uk/PROSPERO/). In the process of implementation, this work will strictly refer to the detailed rules [17] and specifications set in the statement of Preferred Reporting Items for Systematic Reviews and meta-analyses (PRISMA) [18, 21, 22]. This protocol is designed to guide further systematic review work, so there is no need for ethical review. Extracting relevant data from published literature will be the source of data needed in this systematic review, and the process of using literature data does not involve ethical issues.

### Studies of eligibility criteria

#### Types of studies design

We will include randomized controlled trials (RCTs) of acupuncture at SP6 points for PD. The included studies will not consider the restrictions of region, publishing language, publishing status, and publishing year. Non-randomized controlled trials, literature reviews, animal experiments, case reports, medical record discussions, experience summaries, and theoretical discussions will be excluded.

#### Participants

According to the clinical practice guidelines for primary dysmenorrhea issued by SOGC [18, 23], this study will include patients who meet the diagnostic criteria for primary dysmenorrhea. The diagnostic criteria of PD include periodic lower abdominal pain during menstruation or before and after menstruation (within 1 week), sometimes accompanied by soreness of the waist, fatigue, nausea, vomiting, cold limbs, and other discomforts, which affects work and life. After gynecological examination, there is no obvious organic lesion in reproductive organs. This disease mostly occurs in adolescent girls 2 to 3 years after menarche or young women who have not given birth. Regardless of race, nationality, education level, and economic status of patients.

#### Types of interventions

We will include clinical trials of acupuncture at SP6 for PD, excluding those using other methods (acupoint injection, acupoint catgut embedding, acupressure, moxibustion, cupping, transcutaneous electrical nerve stimulation) to stimulate SP6 for PD.

The control group can be SP6 sham acupuncture, placebo, blank group. Studies in which the control group consisted of drug therapy (eg, NSAIDs, Hormonal Treatment, vitamins), acupoint injections, and other complementary alternative therapies (eg, Chinese herbal medicine, yoga, Dietary Supplements) will be excluded. The following treatment comparisons will be considered. (1) SP6 acupuncture / SP6 sham acupuncture; (2) SP6 acupuncture / placebo; (3) SP6 acupuncture / no treatment.

### Outcome measures

#### Primary outcome

The primary outcome will be the Pain index. Visual analogue scale (VAS) to assess changes in pain intensity.

#### Secondary outcome

(1) The recent clinical recovery rate. After treatment, abdominal pain and other symptoms disappeared, and no recurrence occurred in 3 menstrual cycles. (2) indicators of concomitant symptoms. Concomitant symptoms were evaluated by the following scales: Cox menstrual symptom scale (CMSS), menstrual symptom score (MSS), retrospective symptom scale (RSS). (3) Serum prostaglandin. (4) Ultrasonic monitoring of uterine artery blood flow resistance. (5) Adverse reactions, adverse events.

### Literature sources and retrieval strategies

#### General database retrieval and retrieval strategy

Electronic databases will be searched to obtain the required study literature. The upper limit of retrieval time will be set to September 2023. Foreign and Chinese databases will be searched respectively: Cochrane Library, PubMed, EMBASE, Ovid, Korea Med, J-Global, J-Stage databases, and CNKI, CBM, SinoMed, VIP, Wanfang. Randomized controlled trials meeting the inclusion criteria will be screened by two independent reviewers (XW, JY). We will search the above 12 databases using subject terms and free words. The retrieval strategy of the PubMed database is shown in Table 1, and the retrieval strategies of other databases need to be converted accordingly. For the related studies involved in the references, manual retrieval will be adopted.

**Table 1.**
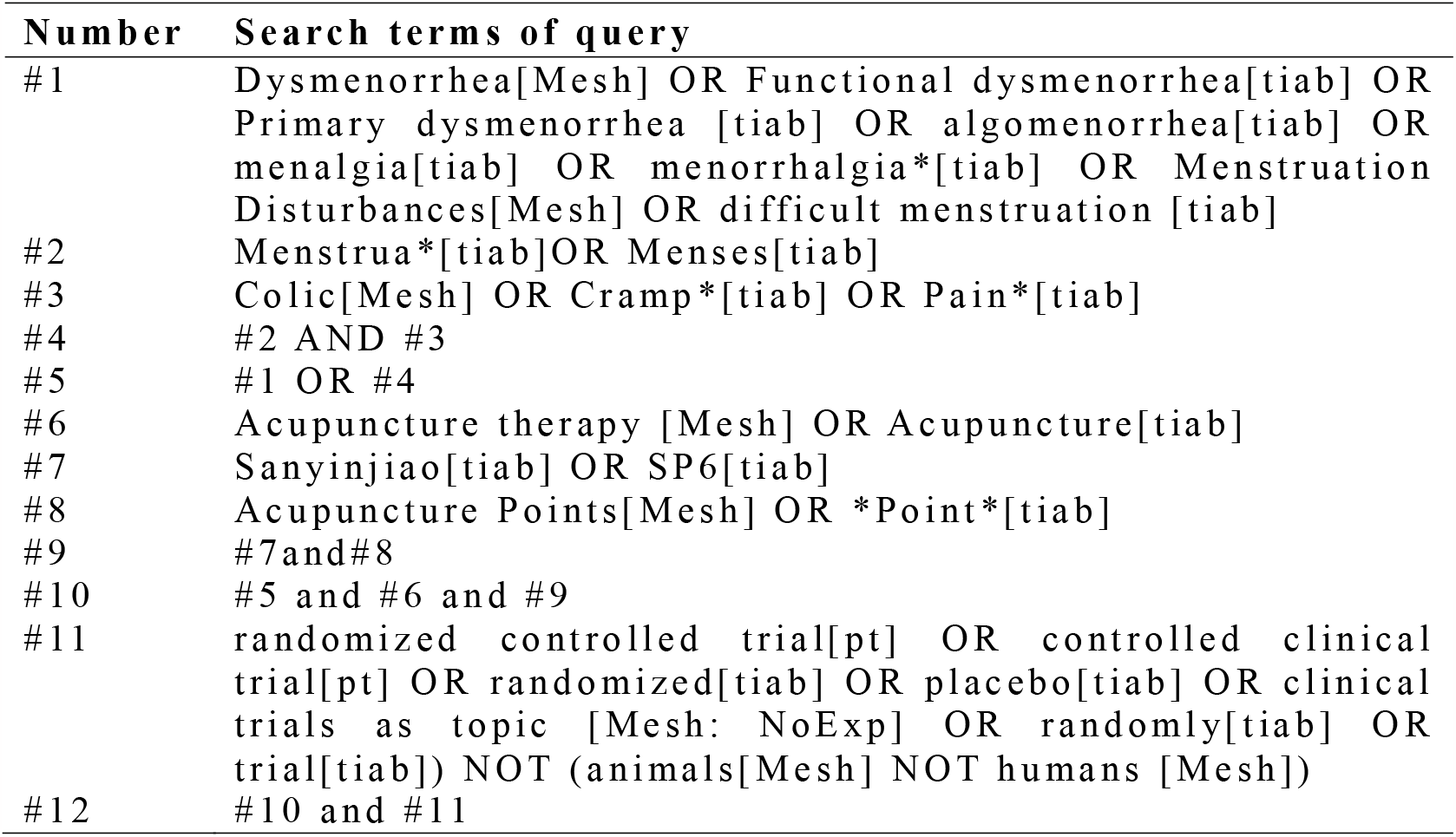
Search strategy for PubMed.

#### Other database resources

To obtain data from ongoing clinical trials, the following databases will be searched: UK National Research Register, WHO ICTRP, ISRCTN, UK Clinical Trial Gateway, Clinic-Trials.gov, UMIN CTR, Chinese Clinical Trial Register. To obtain unpublished clinical trial data, we will continue to search the following databases: OpenGrey, ProQuest, and Conference Literature Database ISI proceeding.

### Data extraction and analysis

#### Study selection

Two independent reviewers (JL, ZZ) will be responsible for the literature screening, and the bibliographies that qualified in the preliminary screening will be imported into the literature management software EndNote (V. X9.0). and then read the titles and abstracts one by one, and eliminate the duplicate or unqualified literature titles. After obtaining the full text of the bibliography, read the full text carefully and evaluate the final included literature according to the inclusion criteria. For the literature that did not meet the inclusion criteria, Excel will be used to record the reasons for exclusion in detail. If the full text is not available in the database, we will contact the corresponding author via email to obtain the full text. If there is a disagreement in the whole process of literature screening, the third reviewer (PX) will help to judge. The process of study screening is shown in the flowchart (Fig 1).

**Figure 1.**
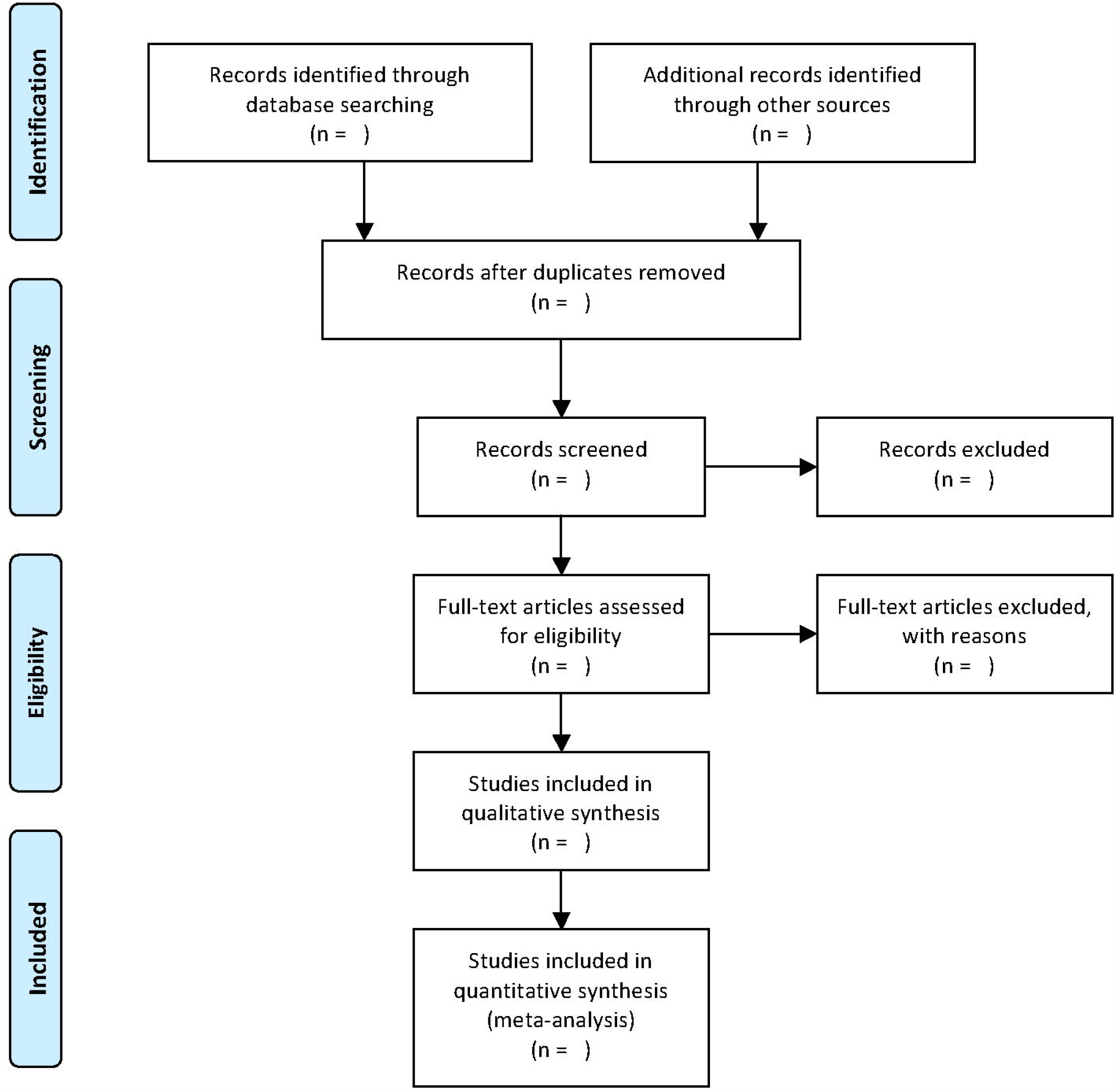
The PRISMA flow diagram of literature screening process. *From:* Moher D, Liberati A, Tetzlaff J, Altman DG, The PRISMA Group (2009). *P*referred *R*eporting *I*tems for *S*ystematic Reviews and *M*eta-*A*nalyses: The PRISMA Statement. PLoS Med 6(7): e1000097. doi:10.1371/journal.pmed1000097

#### Data extraction

The contents of the Excel spreadsheet used for literature data extraction will be decided after discussion by all authors. The contents of the data extraction Excel are composed of five parts: basic information (number, title, author, year, journal, country, fund, language); Study methods (diagnostic criteria, sample size, design type, statistical method, randomized method, treatment plan concealment, blind method, study data integrity, follow-up time); Patient characteristics (course of disease, degree of dysmenorrhea, age, region); Intervention measures (treatment frequency, treatment time, acupuncture group/control group type); Outcome indicators (outcome measurement methods, measurement time points, primary outcome indicators, secondary outcome indicators, efficacy evaluation methods, adverse reactions).

Before the formal literature data extraction, the applicability of the literature extraction form will be tested, and the literature extraction form will be modified according to the preliminary test results to make it more perfect and reliable. Two uniformly trained reviewers (ZZ, JY) will be responsible for formal literature data extraction. If there are differences in the process of data extraction, the two authors will discuss and solve them first. If differences still exist after discussion, the third reviewer (PX) will assist in judging. In the process of data extraction, if the data records are fuzzy and difficult to be extracted, we will contact the corresponding author via e-mail and confirm the data. After data extraction, two reviewers (YY, PX) will cross-check the extraction results and input them into Revman software (V.5.3).

#### Risk of bias assessment

Two reviewers (JY, YY) will carefully read the specific requirements of Cochrane Review Handbook 5.1.0 [18, 19] for bias risk assessment, and will independently use the Cochrane Risk-Of Bias [24] assessment tool to evaluate the risk of bias of the included studies. After the assessment, the two reviewers will cross-check the assessment results. If the assessment results of both parties are different, they will be discussed and resolved through negotiation first. If the differences cannot be resolved after discussion, the third author (XW) shall assist in the judgment. We will evaluate the risk of bias in terms of the method of random sequence generation, whether the allocation scheme is hidden, whether participants are blinded, whether the study outcome evaluation is blinded, the integrity of the outcome data, selective reporting, and so on. Bias risk assessment results are divided into three categories: high risk, low risk, and unclear. The assessment results will be imported into Review Manager 5.3 software to generate the risk of bias graph and risk of bias summary.

#### Measures of treatment effect

If the study results are dichotomous data, we will use odds ratio (OR) and 95% confidence interval (CI) to evaluate the efficacy of acupuncture SP6 in treating PD. If the study results are continuous data, we will evaluate the curative effect of acupuncture SP6 on PD using mean difference (MD), or standardized mean difference (SMD), and 95% confidence interval (CI).

#### Dealing with missing data

If the record of study data is not integrity, we will contact the corresponding author through e-mail, telephone, etc., to obtain complete study data as far as possible. If the complete data is still not available, the research team members will exclude this study after discussion. The impact of missing data on the overall study results will be determined by sensitivity analysis.

#### Assessment of heterogeneity

The heterogeneity among all included studies will be measured by the Chi-square test (χ2) and I^2^ test. If the heterogeneity among the included studies is low, that is, P > 0.1 and I^2^ ≤50%, the fixed-effect model should be used to analyze the study results. If the heterogeneity among the included studies is moderate, that is, P≤0.1 and I^2^ > 50%, the random effect model should be used to analyze the study results. If the heterogeneity among the included studies is high, that is, P ≤ 0.1, I^2^ > 75%, subgroup analysis or descriptive analysis should be conducted.

#### Analysis of publication bias

If the total number of studies finally included in this systematic review exceeds 10, we will use a funnel plot to evaluate publication bias. If necessary, we will further use the Egger regression and Begg correlation test to test the symmetry of the funnel plot. In addition to publication bias, selectivity bias, language bias, and citation bias may also lead to funnel plot asymmetry. If there is an asymmetry in the funnel plot, we will try to analyze the reasons.

#### Data synthesis

The study data will be synthesized and analyzed using RevMan5.3 software according to Cochrane guidelines. If the heterogeneity among the included studies is low, the fixed-effect model should be used to analyze the study results. If the heterogeneity among the included studies is moderate, the random effect model should be used to analyze the study results. If the heterogeneity among the included studies is high, subgroup analysis or descriptive analysis should be conducted.

#### Subgroup analysis

If the analysis of heterogeneity indicates that there is a certain degree of heterogeneity among the studies included in this systematic review, we will use subgroup analysis to determine the source of the heterogeneity. In this study, subgroup analysis can be performed in terms of patient age, depth of acupuncture, frequency of acupuncture, and duration of illness.

#### Sensitivity analysis

The stability of systematic review results can be tested by sensitivity analysis. We will test the stability of meta-analysis results from the aspects of literature quality, sample size, data integrity, and so on.

#### Grading of evidence quality

According to the detailed requirements of the quality of evidence in clinical studies in GRADE guidelines and the classification of the quality of evidence: high, moderate, low, or very low. We will use the GRADE software to evaluate the quality of evidence in clinical studies included in this systematic review.

## Discussion

It is recorded in ancient books of traditional Chinese medicine that SP6 point is good at treating dysmenorrhea, and the clinical application of acupuncture at SP6 in the treatment of PD has gradually increased in clinical practice. Chinese experts agree that acupuncture can be used as an adjunct or alternative to PD therapy [25]. However, 2017 SOGC Clinical Practice Guideline (No. 345-Primary Dysmenorrhea ConsensusGuideline) states that there is insufficient evidence to support the efficacy and safety of acupuncture at SP6 in the treatment of PD, therefore, we conducted a systematic review and meta-analysis to evaluate the efficacy and safety of acupuncture at Sanyinjiao point in the treatment of PD patients. There are inevitable limitations to this systematic review. First, there may be a risk of heterogeneity in the way of acupuncture, the type of needle, and the duration of a single acupuncture treatment, and second, it is difficult to obtain complete raw data from the literature, which may cause reporting bias, which we will clarify in detail. It is hoped that this study will provide the latest evidence analysis on the effectiveness of acupuncture at SP6 in the treatment of PD in order to provide ideas and methods that can be used for reference for doctors, patients, and medical policy-makers.

## Supporting information

Supplemental PRISMA-P 2015 checklist

## Data Availability

After the completion of the systematic review, all the data extracted from the literature will be published with the publication of the article.

## Abbreviations

PD: Primary Dysmenorrhea,
SP6: Sanyinjiao,
VAS: Visual analogue scale,
TCM: Traditional Chinese Medicine.
LOCF: Last Observation Carried forward,
NSAIDs: Nonsteroidal Antiinflammatory Drugs,
WCA: Worst Case Analysis.

## Supporting information

**S1 Checklist** . **PRISMA - P 2 0 1 5 Checklist**

**(DOCX)**

## Author contributions

**Conceptualization :** Xu emei Wan g, PingXie.

**Data curation :** Xu emei Wang, Jiao Yang, Zhiyuan Zhang .

**Formal analysis:** Xuemei Wang, Jing Liu, YepengYang.

**Funding acquisition :** Ping Xie.

**Investigation:** Xuemei Wang, Jiao Yang, Zhiyuan Zhang.

**Methodology :** Xuemei Wang, JingLiu, Yepeng Yang.

**Project administration :** Xuemei Wang, Ping Xie.

**Resources:** Xuemei Wang, Jiao Yang, Zhiyuan Zhang.

**Software:** Xuemei Wang,Jing Liu, Yepeng Yang.

**Supervision:** Xuemei Wang,Jiao Yang, Ping Xie.

**Validation :** Xuemei Wang, Ping Xie.

**Visualization :** Xuemei Wang, Ping Xie.

**Writing – original draft :** Xuemei Wang.

**Writing – review &editing:**Ping Xie.

## Acknowledgements

This work is supported by the National Science Foundation of China (No. 82374512, 81674017).

## Ethics approval and consent to participate

This work is a systematic review and does not need to be submitted to the ethics committee for review.

### Consent for publication

Our research results will be published in peer-reviewed scientific journals, the Prospero website, and relevant academic conferences.

